# “Accumulating harm and waiting for crisis”: Parents’ perspectives of accessing Child and Adolescent Mental Health Services for their autistic child experiencing mental health difficulties

**DOI:** 10.1101/2024.04.09.24305538

**Authors:** Emma Ashworth, Lucy Bray, Claire Hanlon, Harvey Stanway, Georgia Pavlopoulou, David Moore, Bethany Donaghy, Elizabeth Coen, Ellen Firth

**Author notes:** Correspondence concerning this article should be addressed to: Emma Ashworth, Tom Reilly Building, Byrom Street, Liverpool John Moores University, Liverpool, L3 3AF, UK. Contact. **Data Availability Statement** The data that support the findings of this study are available on request from the corresponding author. The data are not publicly available due to privacy or ethical restrictions. **Funding Statement** The authors received no funding for this study. **Ethics Approval Statement** Ethical approval was provided by the authors’ institutional Research Ethics Committee (ref: 23/PSY/046). **Patient Consent Statement** Informed opt-in consent was sought from all participants prior to data collection.

## Abstract

**Background:** Autistic children and young people are at increased risk of mental health difficulties, but often face barriers when seeking help from Child and Adolescent Mental Health Services (CAMHS). There is limited literature exploring the accessibility of CAMHS for autistic young people, particularly from parents’ perspectives. The present study aimed to 1) explore the experiences of parents/carers seeking help from CAMHS for their autistic child’s mental health difficulties, and 2) gain parents’ perceptions of the accessibility of CAMHS support for their child and understand what could be improved.

**Methods:** A mixed-methods survey design was used to learn from parents/carers. 300 parents/carers took part from across the UK between June and October 2023. Quantitative data were analysed using descriptive statistics, and qualitative data using qualitative content analysis.

**Results:** Findings demonstrated the ongoing struggles that parents/carers faced when seeking professional help from CAMHS for their child. Many were not referred to CAMHS or were rejected without an assessment, often due to issues relating to diagnostic overshadowing, a high threshold for assessment, or a lack of professional knowledge about autism and care pathways. Those who were referred reported a lack of reasonable adjustments and offers of ineffective or inappropriate therapies, leaving young people unable to engage, and thus not benefiting. Ultimately, parents felt their child’s mental health difficulties either did not improve or declined to the point of crisis. However, there was a recognition that some professionals were kind and compassionate, and provided the validation that parents needed.

**Conclusions:** There is a need for a more neuro-inclusive and personalised approach in CAMHS, from the professionals themselves, in the adjustments that are offered, and in the therapies that are provided. Further research, funding, and training are urgently needed to ensure mental health support is accessible, timely, and effective for autistic CYP.

## Introduction

Approximately 1% of the UK population are thought to be autistic. Autism is a neuro-biological difference in the way the brain develops before birth and during childhood, meaning that autistic people may process sensory information differently, have different cognitive profiles, and use different communication styles to neurotypical individuals. This influences how they communicate with, behave in, and experience the world. Autistic children and young people (CYP) experience a higher prevalence of co-occurring mental health problems, with around 70% of autistic CYP experiencing one mental health condition and 41% receiving two or more diagnoses, compared to the national average of 12%^1,2^. Similarly, autistic CYP are up to 28 times more likely to think about or attempt suicide than neurotypical CYP^3^ and are more likely to present at Emergency Departments or be admitted to hospital for suicidal crisis^4,5^.

Consequently, autistic CYP are overrepresented in mental health services, with 10% of all CYP using CAMHS thought to be autistic.^6^ Despite this, neurotypical strategies in the assessment of and intervention for autistic CYP’s mental health needs continue to dominate, with disregard for the potential harms of imposing a neurotypical agenda upon them ^7^. Indeed, recent findings^8^ indicated that CAMHS was not considered by parents to be accessible for many autistic CYP, and there are concerns that autistic CYP’s mental health needs are often overlooked in assessments, as symptoms are conflated with traits relating to their autism, a phenomenon known as diagnostic overshadowing^9^. Additionally, symptoms commonly associated with anxiety and depression, such as avoidance, loss of motivation, and sleep disturbance may not be expressed in the same way as in neurotypical individuals, and may be related to autistic-specific experiences such as autistic inertia and/or burnout.^7^ Reports also suggest that CAMHS staff frequently do not feel they have the knowledge or skills to identify and support mental ill health in autistic CYP, and therapists have highlighted concerns over a lack of training.^10^ Thus, autistic CYP experiencing mental ill health are often not assessed or supported effectively, resulting in them reaching crisis.

Recently, multiple national policies focusing on improving mental health outcomes for autistic individuals have been introduced (e.g., NHS Long Term Plan^11^, Transforming CYP mental health^12^, National Strategy for Autistic CYP and Adults^13^, Suicide Prevention Strategy^14^), as well as investments in researching and developing effective interventions. However, access and provision continue to be inequitable, fragmented, and linked to poor outcomes. Service models typically focus on ‘impairments’ or deficits, delivering interventions that respond to difficulties rather than building strengths.^7^ Such neuro-disorder approaches fail to address the impact of a hostile world on autistic wellbeing, and distract staff working with autistic CYP from adopting a neuro-inclusive framework of care.^15^ Ultimately, autistic CYP often do not receive the mental health support they need.^16^

To date, there is limited literature exploring the accessibility of CAMHS for autistic CYP. While studies have examined barriers and facilitators to attaining an autism diagnosis^17^ and accessing psychological support for neurotypical children (e.g.^18^), autistic adults (e.g.,^19^), and autistic children in inpatient settings (e.g.,^20^), there is a paucity of research garnering parents’ perceptions of the accessibility of CAMHS when seeking mental health support for their autistic child. To date, only one study^6^ has examined parents’ perspectives of access to CAMHS for autistic CYP in the UK, and this was over a decade ago. Indeed, a recent meta-analysis of mental health provision for autistic CYP identified a lack of research relating to specialised care pathways, service-wide adaptations, and professionals’ knowledge and skills^9^. Thus, the present study aimed to 1) explore the experiences of parents/carers seeking help from CAMHS for their autistic child’s mental health difficulties, and 2) gain parents’ perceptions of the accessibility of CAMHS support for their child and understand potential improvements.

## Methods

### Design

A mixed-methods concurrent exploratory design^21^ (QUALquan) was used to learn from parents/carers. The qualitative approach was dominant in the study, given the exploratory nature, to allow participants to raise issues and share experiences of importance to them. Ethical approval was provided by the authors’ institutional Research Ethics Committee (ref:23/PSY/046).

### Public and Patient Involvement and Engagement (PPIE)

The team engaged in consultation with autistic adults and CYP, and parents of autistic children, to inform the design, conduct, analysis, and writing up of the study. This is reported according to the GRIPP-2 short form in Table 1.

**Table 1.**
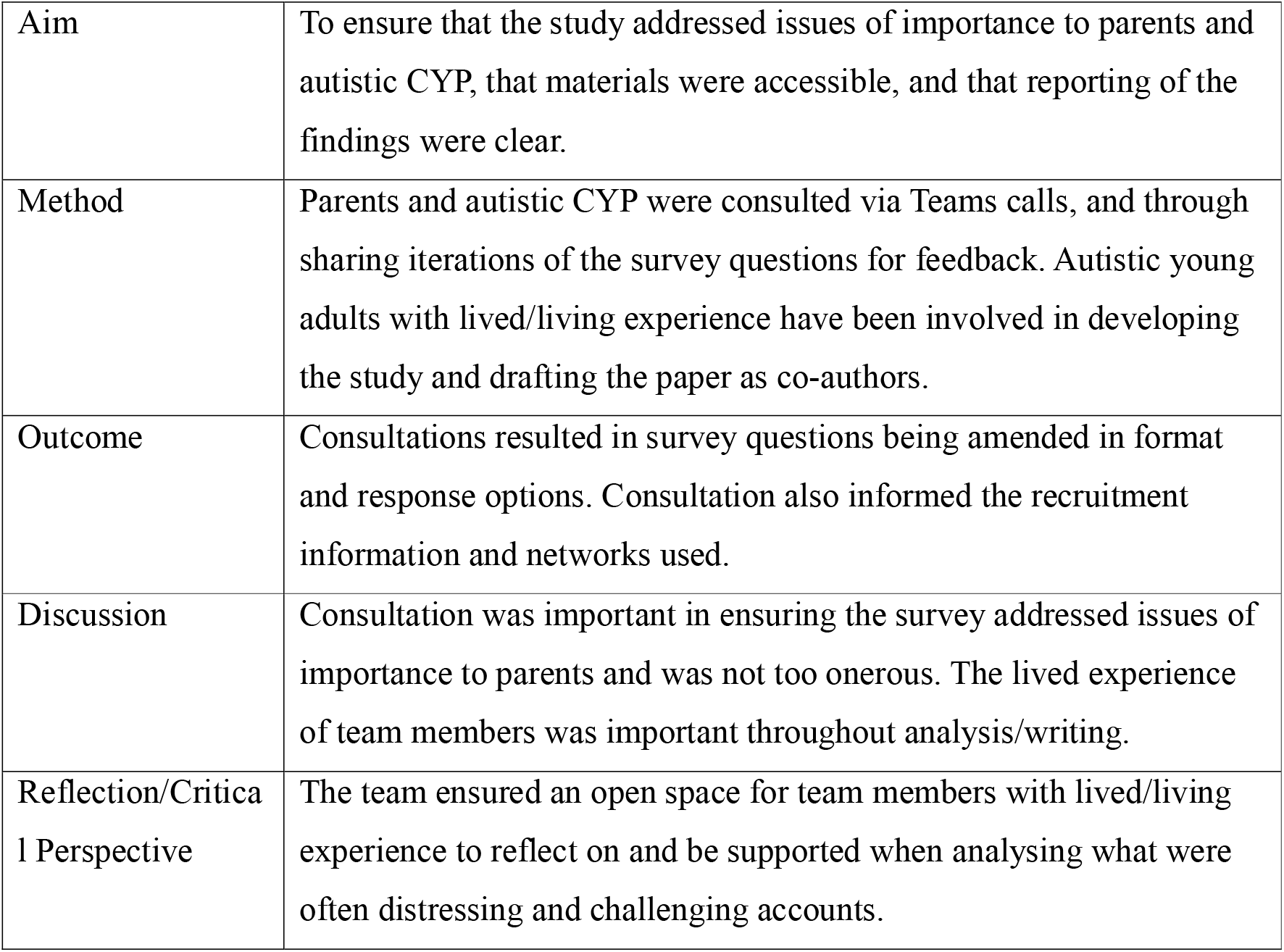
GRIPP-2 Short Form for reporting PPIE.^22^.

### Participants

Parent/carers were recruited using volunteer/opportunity sampling, via social media and through contact with relevant organisations and networks. Parents were invited to take part if they lived in the UK and had an autistic child who had experienced mental health difficulties, for which they had sought professional help in the last five years.

300 parents/carers participated. The majority (87.7%; n=263) lived in England. Their children were aged between 5 and 25 years (mean age=13) and girls were the largest single group (48%; n=144). 84.7% (n=253) also had another long-term health or neurodivergent condition. Table 2 provides further demographic detail. Where percentages do not add up to 100%, there are some responses missing.

**Table 2.**
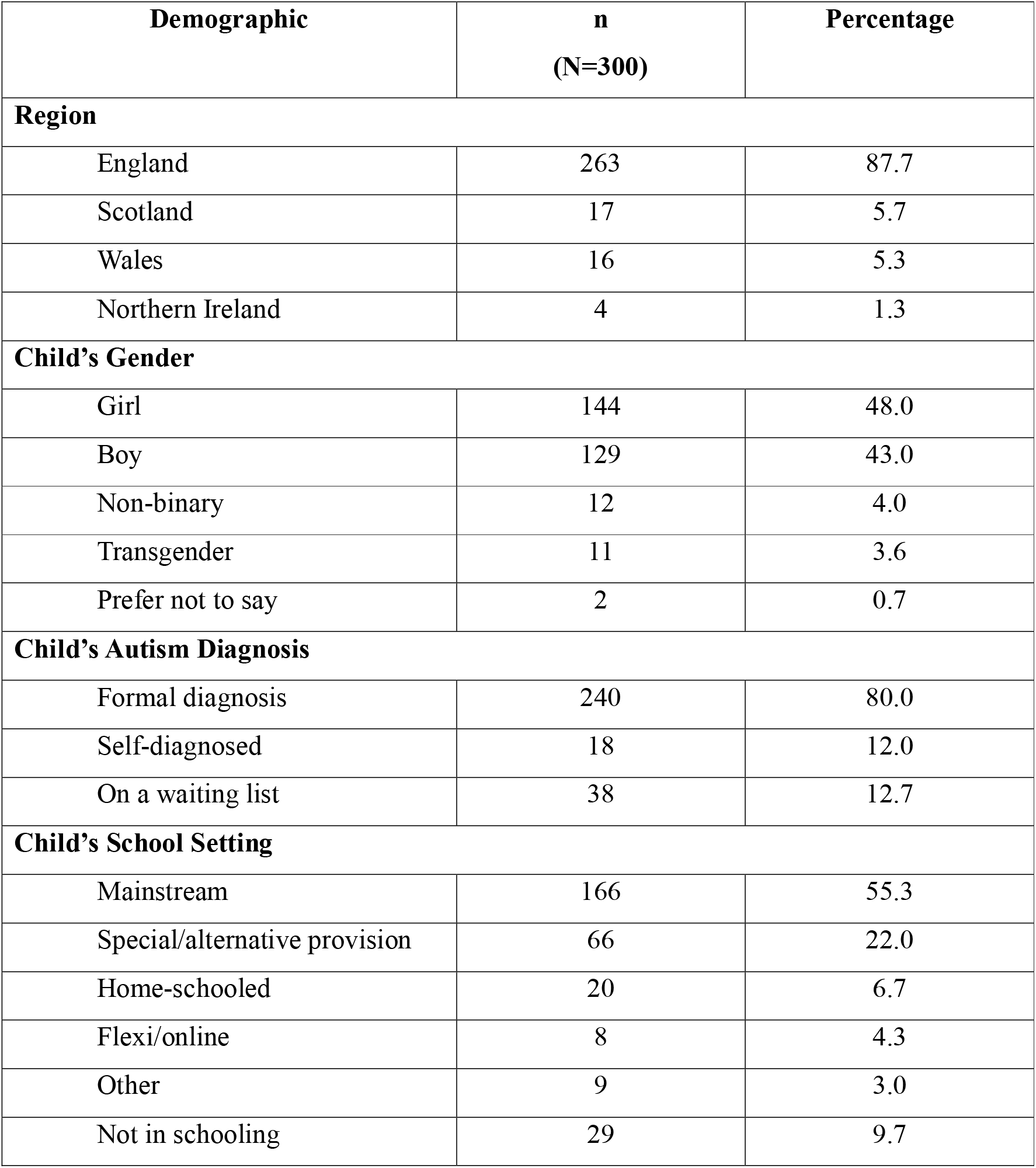
Participant demographic detail.

| **Demographic** | **n**<br>**(N=300)** | **Percentage** |
| --- | --- | --- |
| **Region** |  |  |
| England | 263 | 87.7 |
| Scotland | 17 | 5.7 |
| Wales | 16 | 5.3 |
| Northern Ireland | 4 | 1.3 |
| **Child's Gender** |  |  |
| Girl | 144 | 48.0 |
| Boy | 129 | 43.0 |
| Non-binary | 12 | 4.0 |
| Transgender | 11 | 3.6 |
| Prefer not to say | 2 | 0.7 |
| **Child's Autism Diagnosis** |  |  |
| Formal diagnosis | 240 | 80.0 |
| Self-diagnosed | 18 | 12.0 |
| On a waiting list | 38 | 12.7 |
| **Child's School Setting** |  |  |
| Mainstream | 166 | 55.3 |
| Special/alternative provision | 66 | 22.0 |
| Home-schooled | 20 | 6.7 |
| Flexi/online | 8 | 4.3 |
| Other | 9 | 3.0 |
| Not in schooling | 29 | 9.7 |

### Data Collection

A mixed-methods survey, dominated by exploratory qualitative questions, was developed for this study.^23^ The survey was shared online via QuestionPro (https://eu.questionpro.com/) between June and October 2023. The survey included open- and closed-questions grouped into sections, each exploring a different point along the care pathway (e.g., seeking help, first appointment – see supplementary materials). Surveys began with opt-in consent and demographic questions, before exploring parents/carers experiences of seeking and accessing CAMHS support for their child.

### Analysis

Descriptive statistical analyses of quantitative data were conducted in IBM SPSS v27. Qualitative responses were extracted and collated in Microsoft Word, and were analysed using qualitative content analysis ^24^. Qualitative data were read word-by-word to derive codes, which were subsequently labelled and developed into an initial coding scheme. Codes were then sorted into categories to organise and group them into meaningful clusters. The occurrence of codes was also counted and quantified where appropriate, to understand relative frequency of responses.

## Findings

Findings are presented below, grouped into parents’ experiences at each point of the care pathway explored in the surveys. Relevant quantitative data is presented alongside the qualitative data. As some parents/carers did not answer every question, percentages are based on the total number of responses for each question. Overall, findings portray that, along the care pathway, CYP are reported by their parents/carers as accumulating trauma and experiencing worsening mental health, waiting for inevitable crisis. This summary of the findings, including barriers to accessibility at each point and the resultant impacts, are depicted in Figure 1.

**Figure 1.**
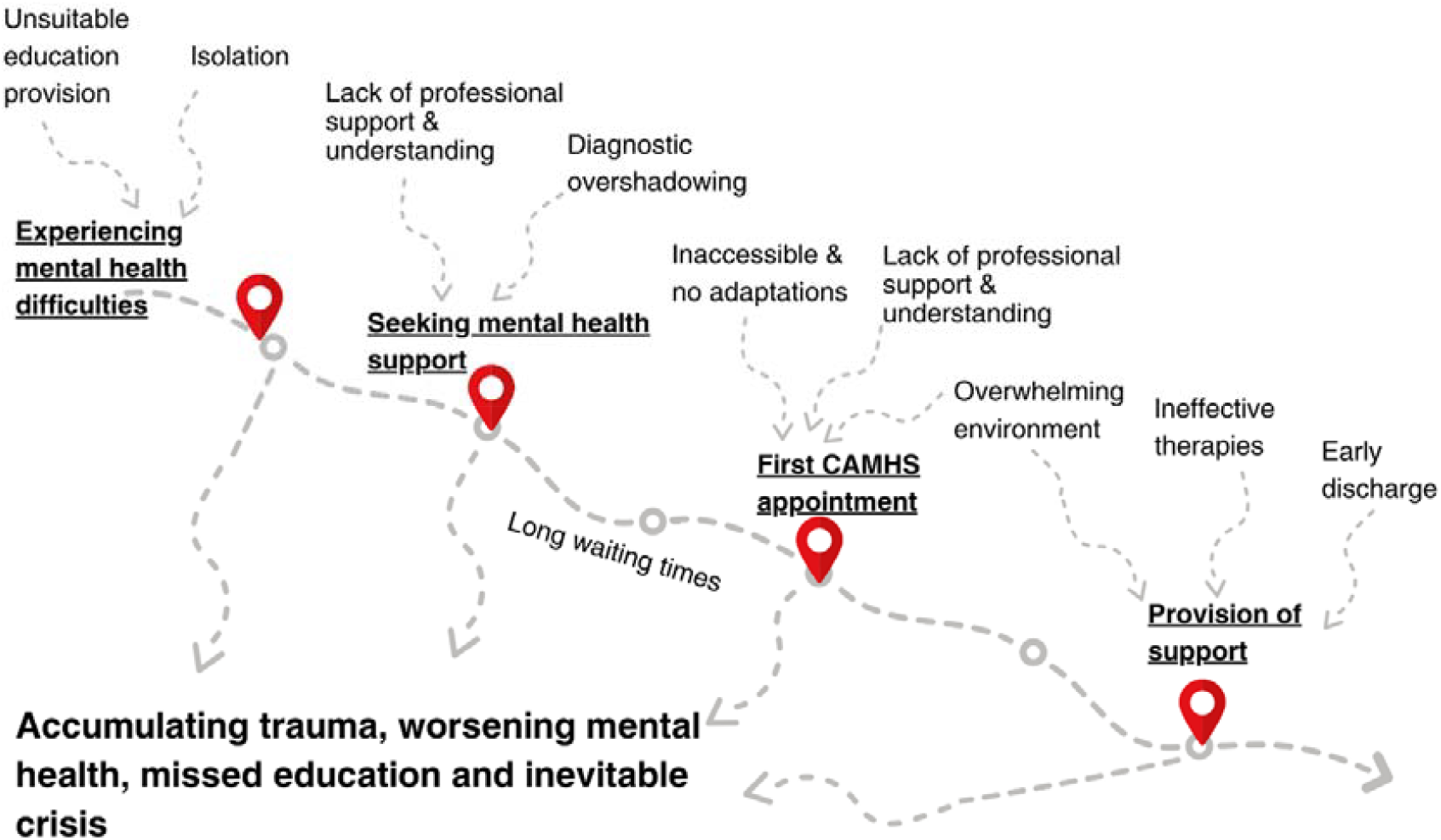
A summary of findings.

### An Overview of Experiences

Figure 2 provides a visual overview of the pathways taken by CYP and their parents when seeking help for mental health difficulties. As can be seen, almost half of those who sought help (46%) were rejected for an assessment by CAMHS, and only a small proportion (9%) of the parents felt that their child’s mental health improved as a result of their interaction with CAMHS.

**Figure 2.**
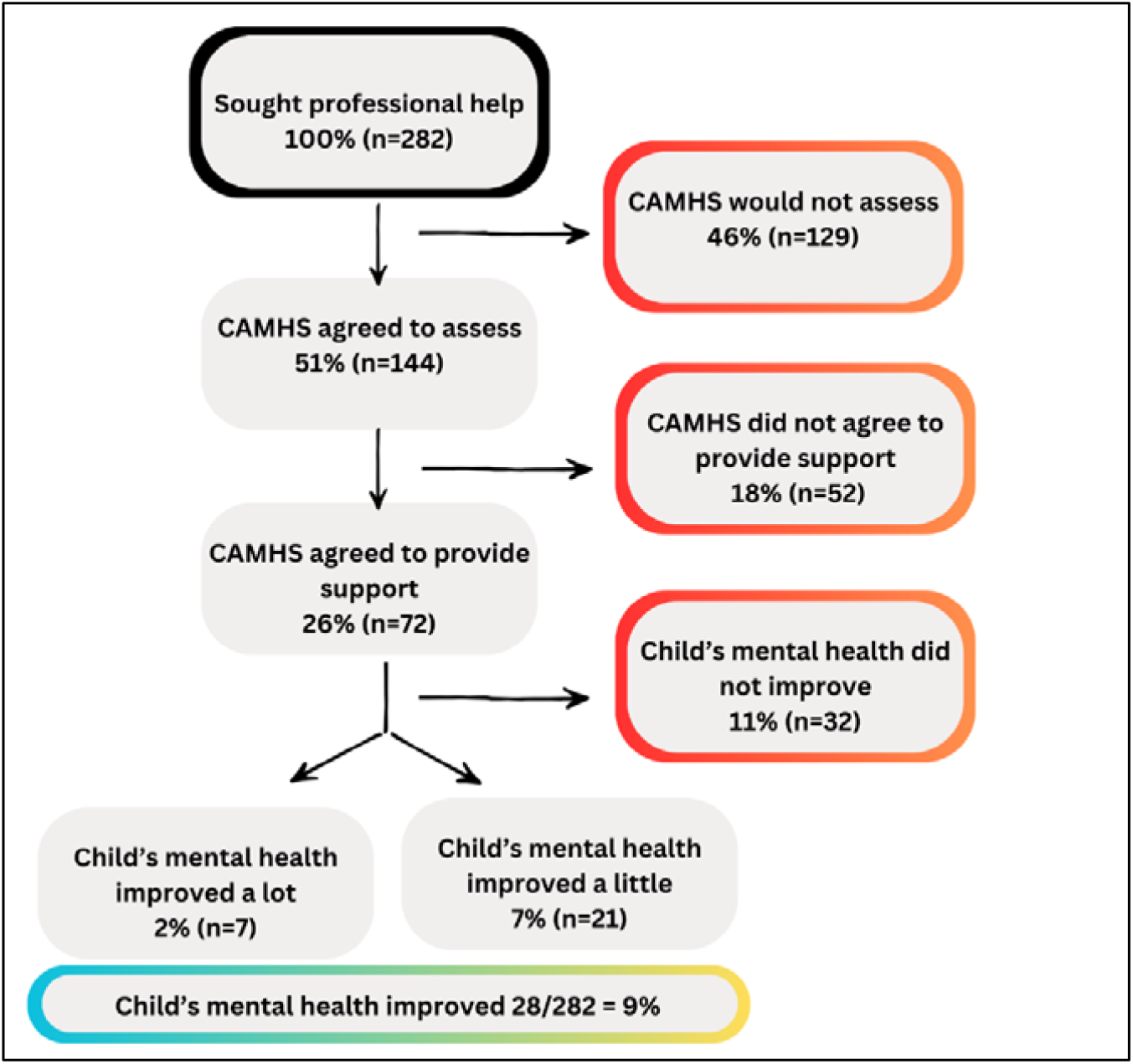
Flowchart of experiences on the care pathway.

### Mental Health Difficulties Leading to Seeking Help

Parents were asked to describe the mental health difficulties that first led to them seeking help for their child. Responses were coded and the number of code occurrences counted. The most frequently cited difficulty was anxiety (n=154), followed by school-based anxiety or difficulties at school (n=96), and suicide ideation (n=83). Table 3 provides an overview of each response type.

**Table 3.**
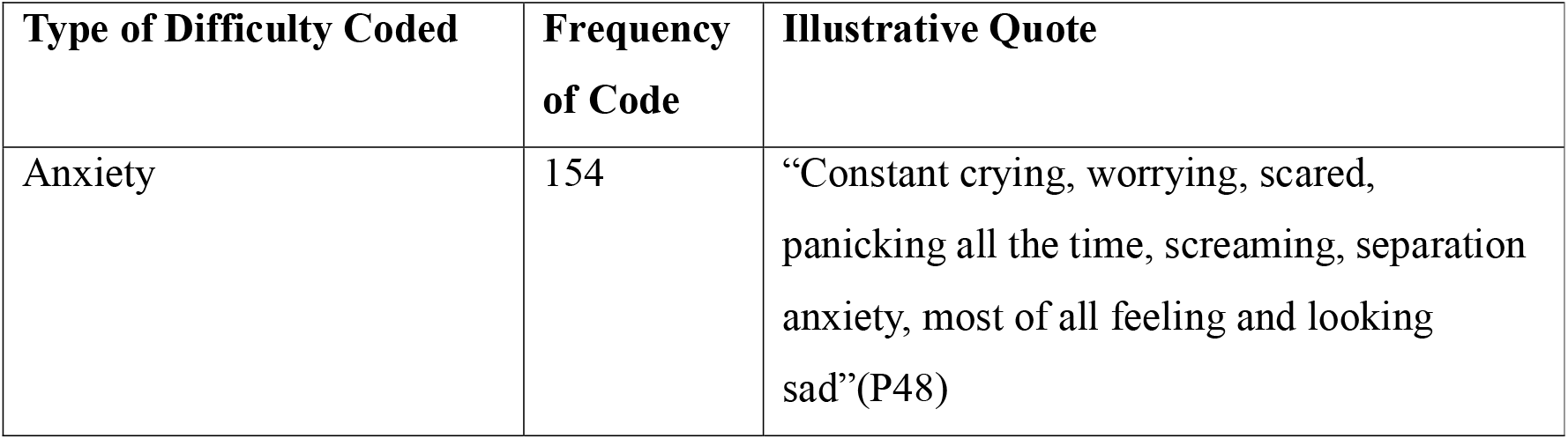
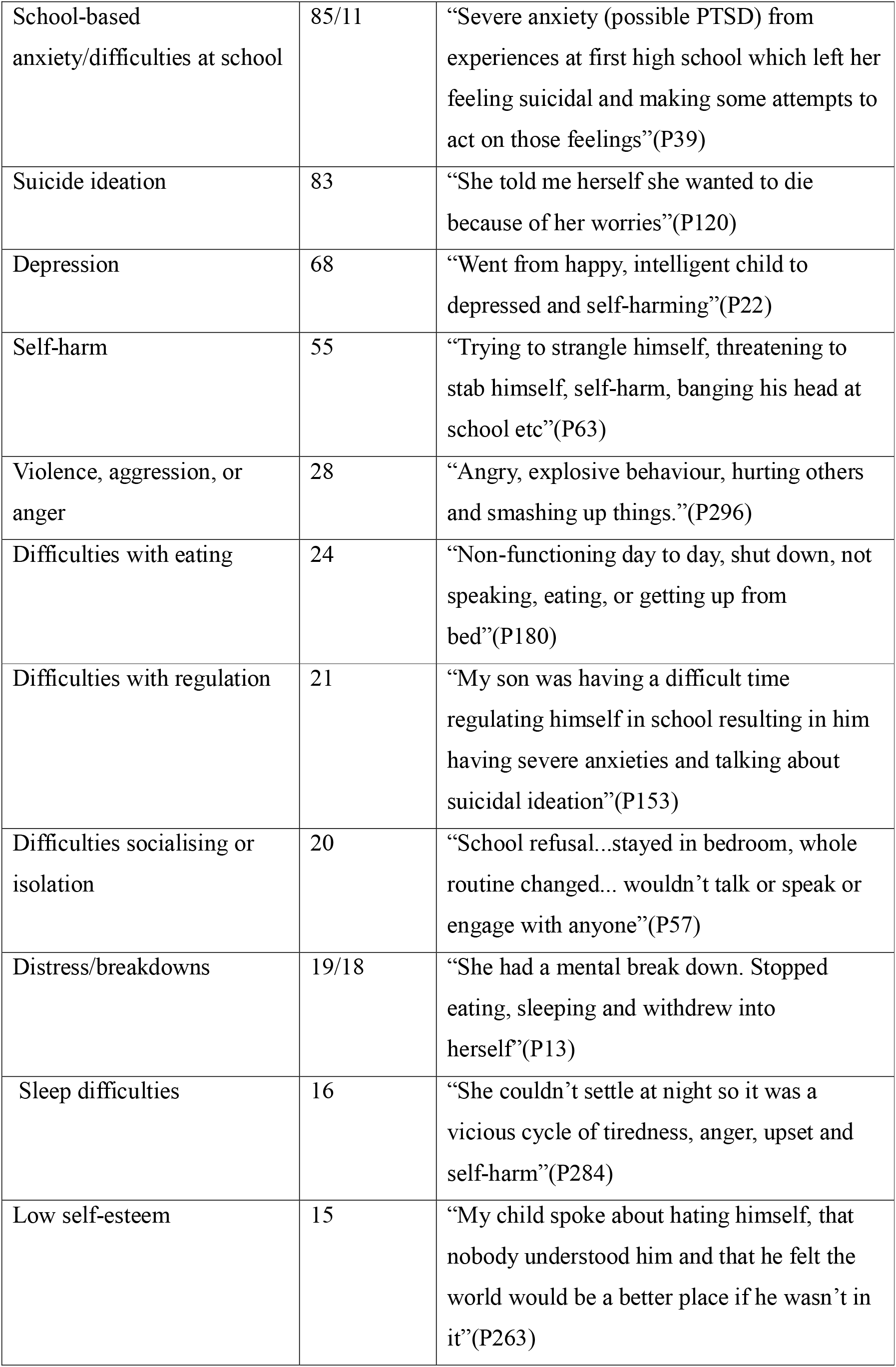
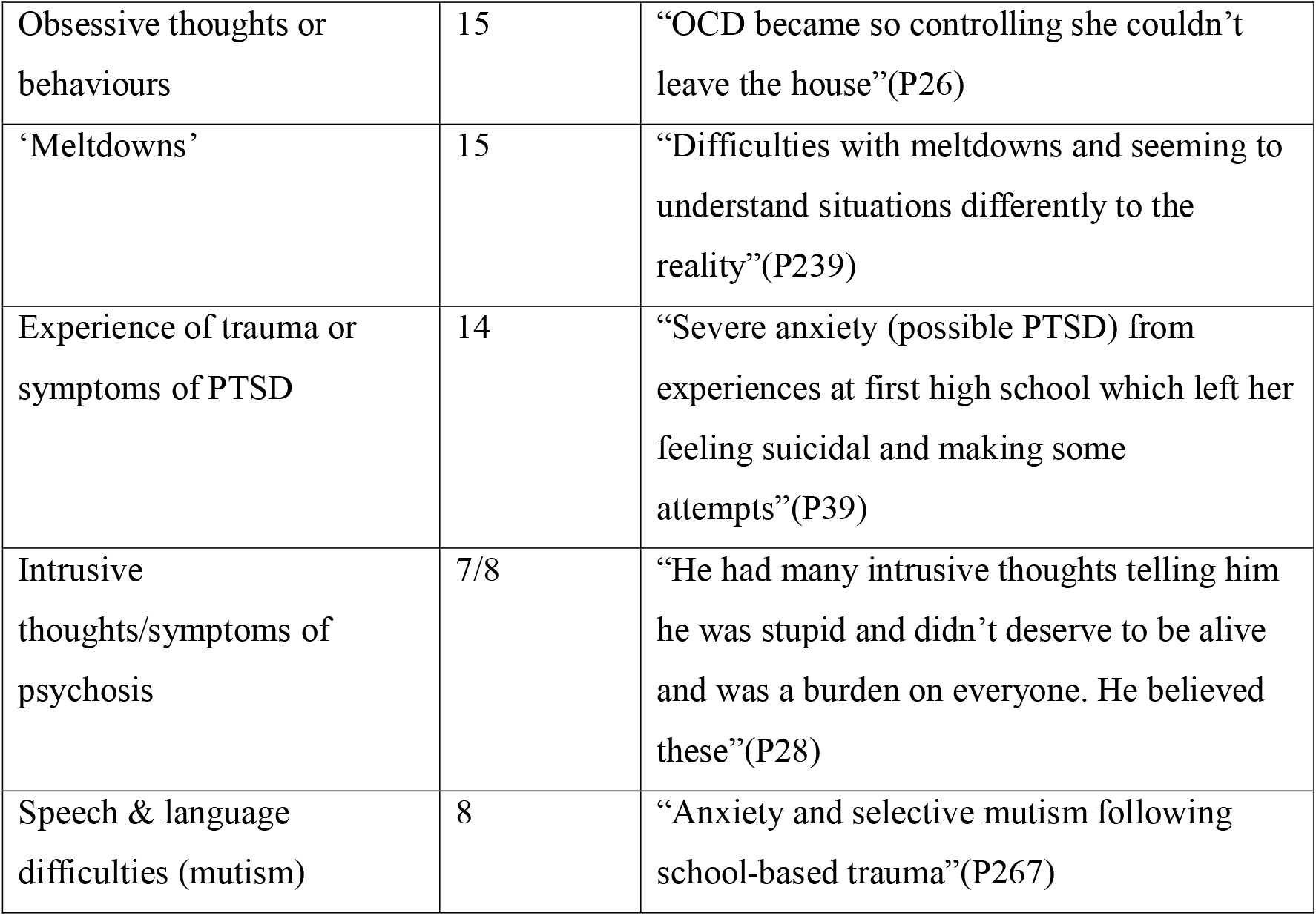
Reasons for seeking help for mental health difficulties.

| Type of Difficulty Coded | Frequency of Code | Illustrative Quote |
| --- | --- | --- |
| Anxiety | 154 | “Constant crying, worrying, scared, panicking all the time, screaming, separation anxiety, most of all feeling and looking sad”(P48) |
| School-based anxiety/difficulties at school | 85/11 | “Severe anxiety (possible PTSD) from experiences at first high school which left her feeling suicidal and making some attempts to act on those feelings”(P39) |
| Suicide ideation | 83 | “She told me herself she wanted to die because of her worries”(P120) |
| Depression | 68 | “Went from happy, intelligent child to depressed and self-harming”(P22) |
| Self-harm | 55 | “Trying to strangle himself, threatening to stab himself, self-harm, banging his head at school etc”(P63) |
| Violence, aggression, or anger | 28 | “Angry, explosive behaviour, hurting others and smashing up things.”(P296) |
| Difficulties with eating | 24 | “Non-functioning day to day, shut down, not speaking, eating, or getting up from bed”(P180) |
| Difficulties with regulation | 21 | “My son was having a difficult time regulating himself in school resulting in him having severe anxieties and talking about suicidal ideation”(P153) |
| Difficulties socialising or isolation | 20 | “School refusal...stayed in bedroom, whole routine changed... wouldn’t talk or speak or engage with anyone”(P57) |
| Distress/breakdowns | 19/18 | “She had a mental break down. Stopped eating, sleeping and withdrew into herself”(P13) |
| Sleep difficulties | 16 | “She couldn’t settle at night so it was a vicious cycle of tiredness, anger, upset and self-harm”(P284) |
| Low self-esteem | 15 | “My child spoke about hating himself, that nobody understood him and that he felt the world would be a better place if he wasn’t in it”(P263) |
| Obsessive thoughts or behaviours | 15 | “OCD became so controlling she couldn’t leave the house”(P26) |
| ‘Meltdowns’ | 15 | “Difficulties with meltdowns and seeming to understand situations differently to the reality”(P239) |
| Experience of trauma or symptoms of PTSD | 14 | “Severe anxiety (possible PTSD) from experiences at first high school which left her feeling suicidal and making some attempts”(P39) |
| Intrusive thoughts/symptoms of psychosis | 7/8 | “He had many intrusive thoughts telling him he was stupid and didn’t deserve to be alive and was a burden on everyone. He believed these”(P28) |
| Speech & language difficulties (mutism) | 8 | “Anxiety and selective mutism following school-based trauma”(P267) |

90% (n=270) of parents reported that their autistic child’s ability to engage with their education had been affected by their mental health difficulties, and many reported that their child’s difficulties were due to attending schools which had outdated perceptions of autism or were unable to meet their needs. As a result, parents reported that their child attended school for an average of 2.7 days per week:

> “The focus is on forcing autistic children to adapt to schools designed for neurotypical children, which is impossible and results in avoidable mental health problems.” (P30)
>
> “[Schools have] an out of date understanding of autism. Better training for teachers and in-school professionals could prevent the need for some autistic children to access CAMHS.” (P194)

Parents’ accounts described the mental health impact of attending an unsuitable education setting: “we wouldn’t have a child with this level of anxiety if they had been able to access education in an environment that made them feel safe, understood, and was flexible to their individual needs” (P62), and the level of anxiety that school caused: “intense anxiety, especially about attending school. Panic attacks, dangerous behaviour such as running into moving traffic to try to avoid going to school.” (P95). One participant described this in more detail:

> “The setting of school can be simply overwhelming to some autistic children (too busy, too many people, too loud, too bright, too many instructions, too many changes etc). School staff are not trained, equipped or resourced to understand how to meet the needs of an ND child.” (P168)

### Seeking Mental Health Support

Parents were asked to indicate who they first spoke to about their child being referred to CAMHS for mental health difficulties. The majority (48.2%; n=136) said they had first approached their GP, followed by school/nursery (27.0%; n=76), a paediatrician or hospital (11.0%; n=31), and 7.8% (n=22) said they self-referred.

56.6% (n=158) felt their child received a referral when they needed it, 41.6% (n=116) felt they did not, and the remainder stated they were not sure. Parents were asked to explain why they did not feel they received the referral when needed, stating issues with diagnostic overshadowing (“it got rejected because ‘anxiety is part of autism’” [P37]), their child not being mentally unwell enough (“only listened to when the self-harm and risk of suicide escalated. We had to be in crisis before we got any help. Kept being told that we didn’t meet threshold” [P38]), long waiting times (“school said they would wait forever, no point” [P41]), poor communication between the school and GP (“doctor said school should refer, school said doctor should refer” [P64]), and schools refusing to refer (“teachers thought there was no issue and my child was just naughty” [P231]).

Of the parents whose child was referred to CAMHS, 50.0% (n=144) said that CAMHS agreed to assess their child, 44.8% (n=129) said they did not, and 8.0% (n=15) were not sure. Where CAMHS did not agree to assess their child, parents were asked to explain the stated reason and what happened next. Common responses included their child not meeting the threshold of need (“I was told she doesn’t meet the criteria as she isn’t in crisis or harming herself” [P103]), diagnostic overshadowing (“they said he’s autistic and they don’t deal with autistic children” [P105]), blaming parenting style (“was told to do parenting course and be stricter” [P94]), that the service was not appropriate for an autistic child (“they felt his ability to engage would not make any programme of work effective enough” [P181]), or that no reason was given (“refused initial referral but didn’t communicate this with anyone!” [P69]). As a result, many were forced to seek private help: “if we had not gone privately, I don’t know where his life would have gone” (P160).

For the children who were added to a waiting list for support, 48.9% (n=68) of parents indicated that their child waited 0-6 months, 18.7% (n=26) waited 7-12 months, 23% (n=32) waited 1-2 years, and 11.5% (n=16) waited more than 2 years. 4.3% (n=6) of parents did not know. 79.1% (n=102) of parents said they were not offered any other NHS support while on the waiting list. For those who were offered support, examples included GP follow-up appointments, parenting courses, and online therapy programmes. Parents described the negative impacts of the long waiting lists without support, highlighting how their children’s mental health continues to worsen to the point of crisis: “you just sit on lists until things get so bad they have to help, or they’re too late and another young person loses their life when it could have been prevented” (P84).

### Attending the First CAMHS Appointment for Assessment

Parents whose child received a referral to CAMHS (N=144) were asked about their experiences of their first appointment for assessment.

66.2% (n=88) reported that their child’s autism diagnosis was disclosed to the CAMHS professional, and 64.0% (n=55) said that this was before the first appointment. Despite this, the majority of parents who had disclosed their child’s autism diagnosis (70.5%; n=55) reported that no reasonable adjustments were made. The majority provided examples of a lack of reasonable adjustments to the environment:

> “Not at all. Waiting room busy and noisy… bright lights and music.” (P111)
>
> “Not in any way which led to the meltdown. Nurse was late. Was left in a busy waiting area and no sensory equipment offered.” (P78)

Of those who indicated that some adjustments were made, many felt these were still ineffective: “they got out some sensory objects but only after she showed signs of distress in the waiting room” (P194). However, a small minority of parents indicated positive adjustments: “lots of sensory toys and fidgets. The CAMHS lady was accepting my daughter wouldn’t look at her and kept jumping into a bean bag whilst talking.” (P256)

53.6% (n=67) reported that their child did not feel at all comfortable explaining their mental health difficulties to the CAMHS professional, 28.8% (n=36) felt ‘a little bit’ comfortable, and 12.8% (n=16) felt ‘quite’ or ‘completely’ comfortable. The remainder did not know or said that their child could not engage. Parents/carers explained that their child did not feel comfortable with the reliance on verbal communication and the expectation for them to be able to understand and articulate their thoughts and feelings to someone they did not know:

> “My daughter struggles to talk to strangers and so I did all the talking away from her while she played” (P184)
>
> “Expecting a 7-year-old to be able to talk about these topics on the phone is a massive ask especially for an anxious autistic child… they should have tried inviting the child into a room with toys/art materials and talked to them while they were playing/creating” (P81)
>
> “She finds it’s difficult to explain what she’s feeling and why” (P74)
>
> “As they are non-verbal it was difficult for them to communicate” (P3)

Many parents explained the negative impact of these often-difficult interactions, as in the following case:

> “Very distressed and overwhelmed… My daughter was forced into a meltdown when the nurse asked around 50 direct questions on the first appointment despite me explaining she wouldn’t cope.” (P78)

While the majority of the experiences were negative, there were a small number of positive comments relating to the kindness of the professionals, which helped to increase comfort: “the person was supportive and kind” (P32) and “as it was the same person each time she built up trust to say what was needed” (P93).

When asked to report how helpful the first CAMHS appointment was, 53.9% (n=69) did not find the appointment helpful for their child, and 24.2% (n=31) said their child could not engage. When asked why they did not find it helpful, parents indicated that they felt blamed for their child’s mental health difficulties: “basically blaming ASD and not parenting my child properly. Really insulting and did not engage well” (P193), or that their child felt blamed, “she was made to feel bad for her worries, questioned a lot, and was embarrassed because she was made to feel silly for her worries” (P120).

Some parents reported that the professionals did not recognise that their child was masking, “they masked, all answers taken at face value when they were fawning” (P137), or have an appropriate level of knowledge about autism: “they didn’t assess him accurately and would not refer him. They did not seem to understand autism. They said his anxiety was because of his autism which is not only untrue, it is a damaging perception of autism.” (P43). Parents also felt that some professionals minimised their child’s difficulties or said they were not severe enough: “CAMHS told me my child had no enduring or significant mental health difficulties. After this appointment my child became very withdrawn and basically went into crisis about 10 days after the appointment.” (P81).

However, similarly to above, there were a small minority of positive comments, which generally focused on the kindness of the professionals, and advice regarding an autism diagnosis: “they were the first professionals who understood and offered reassurance and advice. They also pointed out that she may be autistic.” (P184). Several also reported that they were advised to seek help elsewhere which, while not ideal, did help them ultimately receive the support their child needed: “being told off the record to go privately for therapy.

And luckily we are in the financial position to be able to do this. Most are not.” (P160). Staff within CAMHS were also described by some parents as helping to identify environments which may be harmful to their child’s mental health: “they were the first professionals who actually identified the real problem and solution, which was school and its approach to autism inclusion. Everyone else had blamed our son for his anxiety” (P30).

### The Provision or Offer of Further Support from CAMHS

Parents who received an initial CAMHS appointment (N=144) were asked whether CAMHS agreed to continue to provide support. 55.0% (n=72) reported CAMHS did agree to provide further support, 5.3% (n=7) did not know, and 39.7% (n=52) reported CAMHS did not agree. For the parents who were not offered further support, they were asked why not and where they were signposted to instead. Common responses included that their child was not considered to be engaging enough, did not meet the threshold of need, or were being discharged due to being autistic:

> “Anxiety related to autism and discharged” (P137)
>
> “They told me that she would not be able to access any of their services because she would be too anxious and upset to get to the meetings” (P194)
>
> “Just told I had to call back if got worse or call ambulance if needed” (P89)

Regarding signposting, a large portion of responses stated being directed toward charitable services, as well as informative resources to assist in managing/coping: “was told to read a book” (P197).

The majority of those provided with further support (39.7%; n=27) were offered one-to-one cognitive behavioural therapy (CBT). Other support offered is listed in Table 4. Only 23.4% (n=15) of parents thought that their child felt comfortable or able to partake the support that was offered. When asked to explain their answers, parents typically focused on the need for appropriate, neuroaffirmative therapies which would help their child engage and would not cause further harm or “force the child to mask further” (P41):

**Table 4.** Parents’ reports of support CAMHS offered after the first appointment.

| **CAHMS Support Offered** | **Frequency** | **Percentage** |
| --- | --- | --- |
| CBT (one-to-one) | 27 | 37.5 |
| CBT (group) | 1 | 1.4 |
| Dialectical Behaviour Therapy (group) | 1 | 1.4 |
| Family Therapy | 7 | 9.7 |
| Art Therapy | 2 | 2.8 |
| Other | 20 | 27.8 |
| I was not told | 7 | 9.6 |
| I do not know | 3 | 4.2 |

> “Would like to see access to therapies other than CBT and DBT! I would like to see a service which is neuroaffirmative and seems to understand the needs of autistic young people.” (P300)
>
> “CAMHS causing more harm by offering therapies which are inappropriate e.g. CBT, DBT, PBS – There needs to be safe spaces and trusted and trained adults where they can go to be themselves and therapies to let them express freely, not made to conform.” (P63)

Parents were asked whether they felt their child’s contact with CAMHS improved their mental health. 51.6% (n=32) felt that their child’s mental health did not improve ‘at all’, 33.9% (n=21) felt that it improved ‘a little’, 6.5% (n=4) felt that it improved ‘quite a lot’, and 4.8% (n=3) felt that it improved ‘very much’. Finally, parents were asked what the most positive thing had been; comments mostly focused on the professionals themselves, describing them as “kind people” (P23), and parents explaining “it felt like they wanted to support us” (P188). However, there were very few positive comments generally (“nothing, only medication has kept my son alive” [P286]).

### Improving Mental Health Support

The final questions asked parents what support they would like to see for autistic CYP to help their mental health, both generally and when accessing CAMHS. The majority focused on the benefits of early, preventative interventions to help autistic CYP thrive before mental health difficulties develop: “the length of time or delays in getting support in place means that trauma is inevitable. With effective, efficient scaffolding those ND children are less likely to spiral into mental health crisis” (P168). This early intervention would include improved post-diagnostic support: “there is nothing post-diagnosis which is awful… there should be a separate mental health/autism team who support families once diagnosed!!… we are left with a few agencies who provide short term, like counselling or signposting!!” (P165). Many of the parents also identified the importance of fostering a sense of belonging and community as a protective factor: “we wouldn’t have a child with this level of anxiety if they had met peers who were similar to them… They’d feel less lonely and their anxiety would be reduced. They’d be more independent and have improved self-esteem.” (P62).

Several parents reported that the harm caused to many autistic CYP in accessing education in unsuitable settings could be mitigated by improving staff training, flexible safe spaces, and increased alternative provision: “schools need to be much more aware, flexible and understanding of needs and there have to be autism specialists in every secondary school.” (P241). Several parents also identified the need for interventions to be delivered by dedicated mental health provision for autistic CYP: “an ongoing drop-in service for autistic kids only with practitioners trained in evidence-based therapies proven to be supportive of the autistic neurotype” (P213), and the need for all professionals working with autistic CYP to be up-to-date in neuroaffirmative approaches:

> “CAMHS need to understand what autism means, and use a neuro-affirming approach. They should stop using the basic approach to anxiety which is to assume it is illogical and use exposure therapies. Instead they should investigate whether the anxiety is perfectly logical - for example a stressful sensory environment.” (P80)
>
> “An understanding that autistic children present their mental health problems differently and are as entitled to universal, targeted and specialist mental health support… Then support, advise, assessment and interventions that are neurodevelopmental trauma-informed, autism-appropriate, and neuroaffirmative.” (P37)

## Discussion

This mixed-methods study addresses gaps in understanding around the experiences of parents seeking help from CAMHS for their autistic child. Findings highlight the health inequalities experienced by this group of CYP, who are already at increased risk of experiencing mental ill health and suicidality. A summary of the issues identified, along with associated considerations and recommendations for practice, are provided in Table 5; however, key points are also discussed below.

**Table 5.**
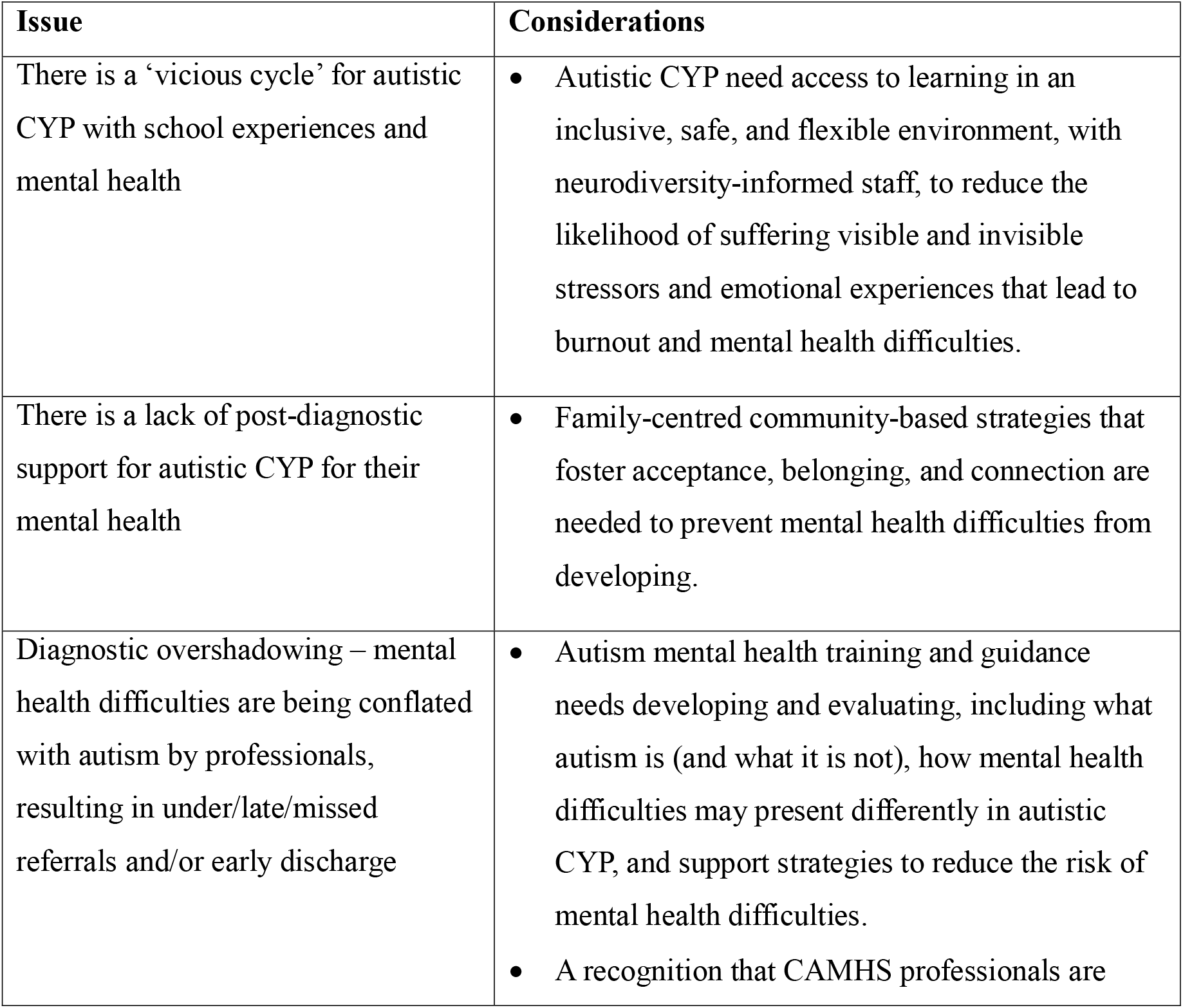
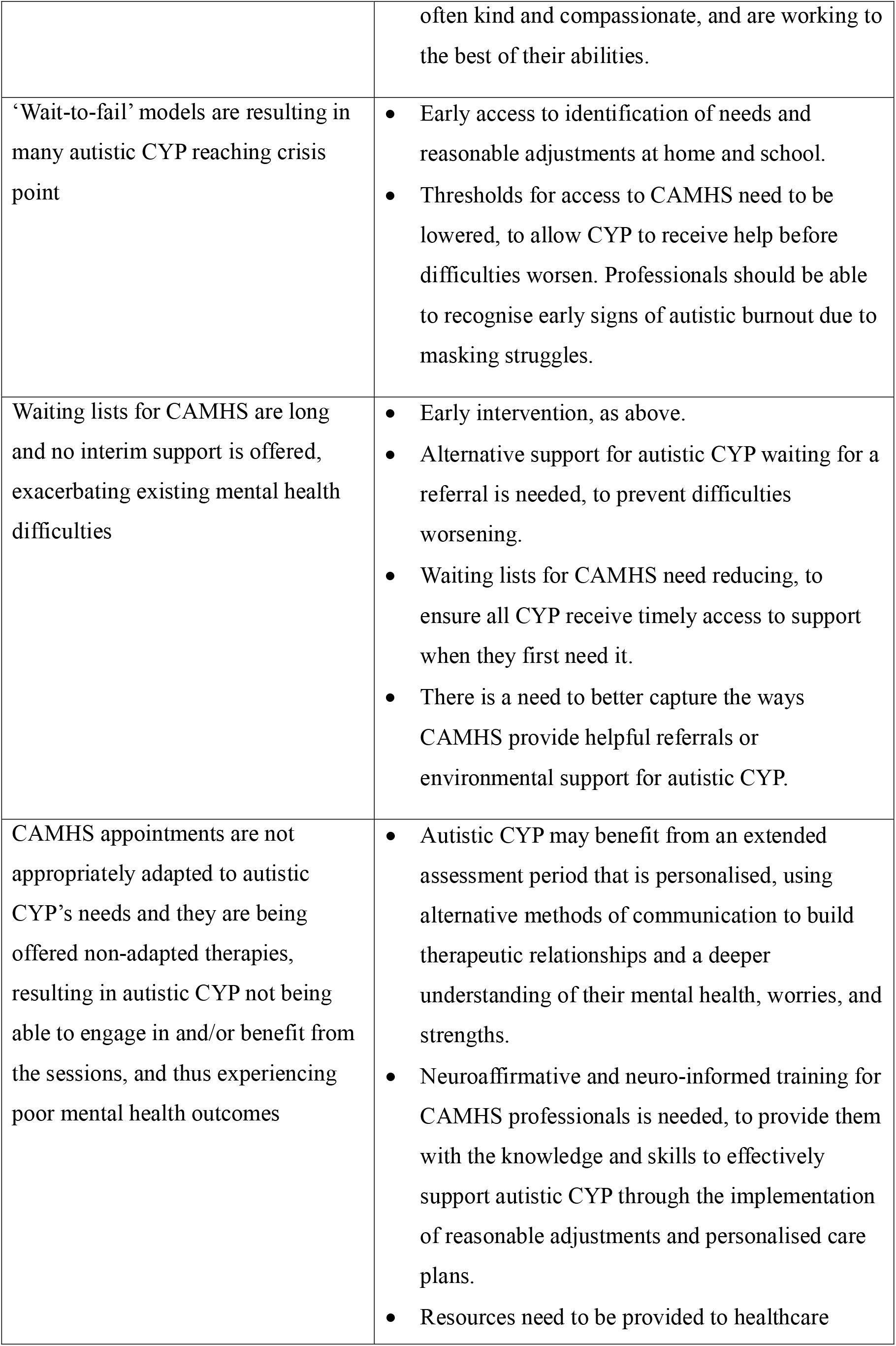
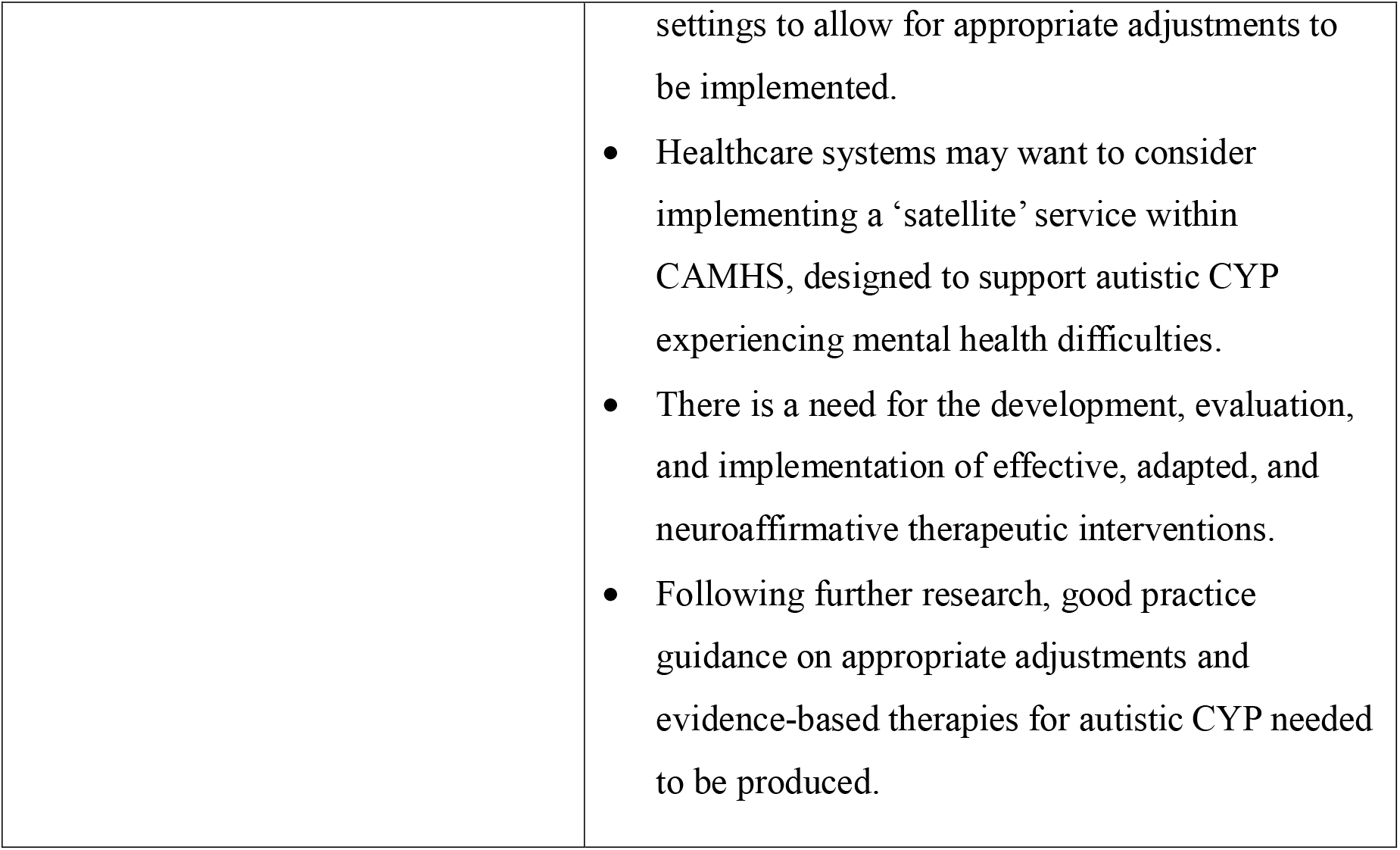
Summary of key issues and associated recommendations.

| Issue | Considerations |
| --- | --- |
| There is a 'vicious cycle' for autistic CYP with school experiences and mental health | Autistic CYP need access to learning in an inclusive, safe, and flexible environment, with neurodiversity-informed staff, to reduce the likelihood of suffering visible and invisible stressors and emotional experiences that lead to burnout and mental health difficulties. |
| There is a lack of post-diagnostic support for autistic CYP for their mental health | Family-centred community-based strategies that foster acceptance, belonging, and connection are needed to prevent mental health difficulties from developing. |
| Diagnostic overshadowing – mental health difficulties are being conflated with autism by professionals, resulting in under/late/missed referrals and/or early discharge | Autism mental health training and guidance needs developing and evaluating, including what autism is (and what it is not), how mental health difficulties may present differently in autistic CYP, and support strategies to reduce the risk of mental health difficulties.<br>A recognition that CAMHS professionals are |
|  | often kind and compassionate, and are working to the best of their abilities. |
| ‘Wait-to-fail’ models are resulting in many autistic CYP reaching crisis point | • Early access to identification of needs and reasonable adjustments at home and school.<br>• Thresholds for access to CAMHS need to be lowered, to allow CYP to receive help before difficulties worsen. Professionals should be able to recognise early signs of autistic burnout due to masking struggles. |
| Waiting lists for CAMHS are long and no interim support is offered, exacerbating existing mental health difficulties | • Early intervention, as above.<br>• Alternative support for autistic CYP waiting for a referral is needed, to prevent difficulties worsening.<br>• Waiting lists for CAMHS need reducing, to ensure all CYP receive timely access to support when they first need it.<br>• There is a need to better capture the ways CAMHS provide helpful referrals or environmental support for autistic CYP. |
| CAMHS appointments are not appropriately adapted to autistic CYP’s needs and they are being offered non-adapted therapies, resulting in autistic CYP not being able to engage in and/or benefit from the sessions, and thus experiencing poor mental health outcomes | • Autistic CYP may benefit from an extended assessment period that is personalised, using alternative methods of communication to build therapeutic relationships and a deeper understanding of their mental health, worries, and strengths.<br>• Neuroaffirmative and neuro-informed training for CAMHS professionals is needed, to provide them with the knowledge and skills to effectively support autistic CYP through the implementation of reasonable adjustments and personalised care plans.<br>• Resources need to be provided to healthcare |
|  | settings to allow for appropriate adjustments to be implemented.<br>• Healthcare systems may want to consider implementing a ‘satellite’ service within CAMHS, designed to support autistic CYP experiencing mental health difficulties.<br>• There is a need for the development, evaluation, and implementation of effective, adapted, and neuroaffirmative therapeutic interventions.<br>• Following further research, good practice guidance on appropriate adjustments and evidence-based therapies for autistic CYP needed to be produced. |

Findings demonstrated the ongoing struggles that parents faced when seeking help from CAMHS. Many of their children were either not referred to CAMHS to begin with or were rejected without an assessment, often due to issues relating to diagnostic overshadowing, a high threshold for assessment, or a lack of professional knowledge about autism and care pathways. Those who were referred reported a lack of reasonable adjustments and offers of ineffective therapies, leaving CYP feeling uncomfortable, unable to engage, and thus not benefiting from the service. Ultimately, parents felt their child’s mental health difficulties either did not improve or declined to the point of crisis, felt blamed by clinicians, and reported an overall lack of support. Consequently, parents expressed a need for a more neuroaffirmative approach in CAMHS, from the professionals themselves, in the adjustments offered, and in the therapies provided. Worryingly, the findings closely align with those from Read and Scofield^6^ conducted almost fifteen years ago, highlighting that little has changed regarding access to CAMHS for autistic CYP. Similarly to findings presented here, they reported that the majority of parents felt that CAMHS had not improved their child’s mental health and that they had encountered difficulty in accessing CAMHS. They also reported a lack of professional understanding for their child’s autism, resulting in what felt like inappropriate, ineffective, and even harmful therapies.

While little other evidence from parents’ or children’s perspectives exist, a recent report from ‘Spectrum Gaming’, a charity for autistic CYP,^25^ presented the perceptions of autistic CYP who had previously accessed CAMHS for mental health difficulties. They suggested that autistic CYP felt the support they received often centred around identifying symptoms and deficits rather than addressing the root causes of problems, that anxiety and autism are considered by professionals to be synonymous, that CAMHS should offer appropriate and effective therapies, and that needs to be a greater understanding of autism within services. Additional evidence exists highlighting how parents feel blamed by professionals for their autistic child’s mental health difficulties.^26^ More generally, a systematic review of barriers and facilitators of healthcare access for autistic CYP^27^ identified many similar issues, including a lack of professional knowledge about autism, sensory issues preventing engagement, and system-level barriers including a lack of resources for adjustments, long waiting times, and a lack of person-centred care. Furthermore, it appears this issue is not specific to CYP, with autistic adults also reporting that they do not feel adequately supported by mental health services and that some treatments are ineffective or cause additional harm, calling for a flexible and holistic approach.^19^

Results from the current study highlight clear implications for policy and practice, as well as future research directions, in order to better meet the needs of autistic CYP in CAMHS and reduce the likelihood of further harm. Findings from the current study suggest that CAMHS professionals may want to help and be willing to implement support, but may be limited by a lack of knowledge and resources. Indeed, one of the only clear positives highlighted by parents when accessing CAMHS was the kindness, compassion, and validation provided by some staff. Thus, there is a need for further training amongst professionals, and an evaluation of that training, to ensure all have a clear understanding of what autism is (and what it is not), how mental health difficulties might present in autistic CYP, and reasonable adjustments that could be implemented. This training should be developed with autistic CYP, ensuring their voices are heard regarding what works for them. However, in order for professionals to make these adjustments, resources and funding need to be provided for their implementation. This particularly pertinent given the current NHS Long Term Plan,^11^ which outlines the NHS’s plans to improve its understanding of the needs of autistic people, including providing healthcare staff with training on supporting autistic people and ensuring they make reasonable adjustments.

In addition, to be able to meaningfully reduce mental health difficulties in autistic CYP, further research is needed into the appropriateness and effectiveness of therapies, as there is currently little robust evidence regarding the utility of adapted/alternative therapies for autistic populations. This information could be used to inform the development of guidelines for clinicians; The National Autistic Society^28^ recently shared a practical guide which may be a useful starting point. Finally, there is a wider issue within CAMHS generally, whereby thresholds for assessment are too high and waiting lists are too long, resulting in a reactive ‘wait-to-fail’ model^29^ and CYP reaching crisis point.^30^ While this is not specific to autistic CYP, it can disproportionately negatively impact this group who are already vulnerable to health inequalities. Thus, there is a need for more funding and service improvements to enable earlier intervention, rather than waiting for difficulties to escalate.^31^

Finally, a ‘vicious cycle’ between schooling and mental health was frequent for autistic CYP. Many parents felt that school caused their child’s mental health difficulties to begin with, and that this was compounded over time. For many parents, their child’s mental health difficulties then meant they struggled to attend school, resulting in their educational progression and achievement also being negatively impacted. Indeed, parents reported that, on average, their child was only attending school for half of the amount of time that they should be. There is thus a clear need for similar work in education settings, promoting early intervention through the provision of resources for adaptations, staff training, and evidence-based interventions.

### Limitations

The current study has several limitations that need to be considered. Firstly, parents/carers in this study self-selected and, as such, may not be representative of the entire population who have sought help from CAMHS. Indeed, the parents who had a more positive experience may not have felt the need to participate. Furthermore, as the survey was online, some parents/carers may not have had access to the necessary technology; thus, it is possible that the families who are most isolated were not reached. In addition, this study only captured the views of parents/carers, and not CYP themselves, meaning that some vital experiences may have been missed. Future research should seek to explore this.

## Conclusion

Findings from this study highlight the ongoing struggles that parents face when seeking help from CAMHS for their autistic child’s mental health difficulties. Many children were not able to access CAMHS due to issues relating to diagnostic overshadowing, a high threshold for assessment, long waiting lists, and a lack of professional knowledge about autism. While CAMHS is already facing difficulties meeting demand generally, these issues are amplified for autistic CYP, a group who face disproportionate health inequalities and are more likely to experience mental ill health. Parents expressed a need for a more neuroaffirmative approach in CAMHS, as well as more effective and informed support from schools. Further research, funding, and training are urgently needed to ensure mental health support is accessible, timely, and effective for autistic CYP.

## Data Availability

The data that support the findings of this study are available on request from the corresponding author. The data are not publicly available due to privacy or ethical restrictions.

## Acknowledgements

We would like to thank all of the parents/carers who have shared their views, experiences, and time with us. We would also like to thank the parent advisors and young people we consulted to help design the study.

